# Reproduction number estimation using smoothing methods Reproduction number and smoothing methods

**DOI:** 10.1101/2025.07.05.25330940

**Authors:** Mahamadou Barro, Serge Manituo Aymar Somda, Cédric Stephane Bationo, Bernard Eric Agodio Dabone, Betty Kazanga, Fabrice Anyirekun Some, Benjamin Roche, Jean Gaudart

## Abstract

The reproduction number for an epidemic is crucial as it helps to understand the potential spread of an infectious disease within a population. It is often estimated through case counting and contact tracing methods. Here, we propose an alternative approach based on real-time smoothing techniques. A generalized additive model (GAM) is proposed to model the number of new infectious cases. The characteristics of the smoothing functions are used to estimate the basic reproduction number, R_0_, and the effective reproduction number, R_t_. The proposed method is assessed on simulated data. We simulated three epidemic scenarios with different transmissibility levels, corresponding to different R_0_ values (11.52, 2.88, and 1.06), using a stochastic SEIR model. The method was then compared with validated methods already implemented in R software: the exponential growth method developed by Obadia et *al*. and the renewal process method developed by Cori et *al*. Our splines smoothing method estimated credible tolerance intervals and provided a better fit to data variations while maintaining an appropriate balance between bias, precision and coverage. This reflects the method’s flexibility and ability to capture the epidemic dynamics. In the three scenarios, the splines smoothing method produced median R_0_ estimates of 12.33 [8.34-18.03], 2.33 [1.56-3.56], and 1.31 [0.77-3.45]. The renewal process method led to credible median R_0_ estimates of 12.78 [10.48-17.48], 2.51 [2.02-5.29] and 1.91 [1.23-3.98], respectively. Neither method exhibits a distinct superiority across all contexts. Their efficacy varies depending on performance priorities. In contrast, the exponential method showed inferior performance relative to the other two approaches, exhibiting moderate bias and potentially lower coverage rates with median R_0_ estimates of 13.90 [10.39-33.47], 2.29 [1.08-7.6] and 1.07 [0.39-5.56]. Real-time smoothing provides results comparable to established classical methods while retaining the full flexibility of generalized additive models.

**Author summary:** In the context of infectious disease outbreaks, understanding the spread rate is crucial. The reproduction number, which represents the average number of secondary infections generated by an infected individual is a widely used metric. This number can be estimated at the onset of the epidemic (basic reproduction number, R_0_) or at a specific point in time (effective reproduction number, Rt). The objective of this work is to propose an estimate based on statistical smoothing methods, specifically the Generalized Additive Model (GAM) smoothing method. This method provides a more accurate estimate of the epidemic’s initial phase and estimates R_0_ as effectively. The strength of the splines smoothing method lies in its adaptability across different, scenarios, allowing it to track changes in the data. We compared this approach to two commonly used methods for estimating the reproduction number. While the exponential growth method is simple but less efficient, the spline smoothing method demonstrated comparable efficacy to the renewal process method. Furthermore, it identifies the initial phase of the epidemic with rigor and simplicity and y and offers flexibility by allowing various smoothing techniques.

## 1. Introduction

The SARS-CoV-2 pandemic (2019-2023) [1] has underscored the inherent challenges in risk assessment and management, with significant socio-economic consequences. The primary goal of epidemic anticipation is to implement interventions that will mitigate the impact of the outbreaks, including economic and social effects. For over a century, researchers have utilized various mathematical models to understand epidemic transmission factors [2–6]. These models could be deterministic,, agent-based, stochastic, network, metapopulation, or statistical. Nevertheless, many of these rely on the individual classification through compartment, such as the SEIR (Susceptible-Exposed-Infectious-Removed) models.

The principal indicator for evaluating the spread of an epidemic is the reproduction number, which can be either basic (R_0_) or effective R_t_. These indicators represent the mean number of new (secondary) infections caused by an infected and contagious individual either at the start of an epidemic or at any given time point (t). Several studies have sought to estimate the number of basic reproductions of the SARS-CoV-2 virus worldwide [2–6]. Numerous methods for estimating the reproduction number have been documented. However, the application of mechanistic methods to transmission [5,7–9] is limited by specific assumptions tied to the context, complicating their generalizability. Similarly, statistical methodologies [10–14], may produce estimates that vary accordingly on the selected time interval.

The current approaches rely on the assumption that the case counts from surveillance data are both complete and accurate. Effective epidemic response depends on a well-informed and efficiently managed surveillance system for early detection of cases, intervention planning, and impact evaluation. Such data support the real-time adjustment to strategies based on epidemic trends and facilitate the coordination of efforts among various stakeholders involved.

Here, we present a method that fits a model to available data to estimate the reproduction number, suggesting a capacity to account, to some extent, for observation bias. The objective of our method was to estimate R_t_ each time and R_0_ using smoothing methods based on generalized additive models (GAM). The distinctive feature of this method was its ability to not only estimate the reproduction number but also to assess the initial epidemic phase, essential for estimating R_0_, in a straightforward manner.

## 2. Materials and methods

### 2.1. Methods for Estimating the reproduction number

#### 2.1.1. The basic reproduction number and the effective reproduction number

There are several methods used to estimate reproduction number (for further details, see [16–27]). Over the last decade, significant advancements have been made in statistical methods for estimating reproduction numbers using surveillance data [10–13,15,29], starting with the pioneering work of Wallinga and Teunis [28] and later by White et *al*. [21].

A recent work, inspired by the work of White et *al*. [10,21], has been specifically designed for joint estimates of the effective reproduction number and the serial interval without relying on additional contact tracing data. The likelihood function L(R_t_, p|N) for R_t_ and p (the serial interval distribution) is constructed on the assumption that the total number of cases reported each day follows a Poisson distribution, with subsequent infections following a multinomial distribution.

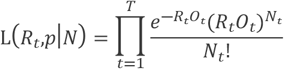

where

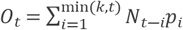

The R_t_ value is represented as a function of time using an interpolation technique, namely the Adaptive Weighted Neighbour (AWN) method, with the objective of capturing the fluctuations observed throughout the course of the epidemic.

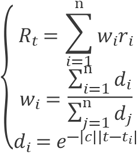

with *t*_*i*_ ≥ 0, *r*_*i*_ ≥0, *n* ≥ 2, *i* = 1,2,…,*n*

The Robust Adaptive Metropolis (RAM) algorithm is applied to perform an MCMC procedure for the estimation of the target distribution of R_t_ and p.

#### 2.1.2. Estimation using the Poisson regression by Obadia et al. [30]

The exponential growth (EG) hypothesis, as developed by Wallinga and Lipsitch [27], serves a foundational method in this context. In this method, the number of new cases, I(t), at time t is modelled as following an exponential distribution:

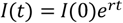

where r is the exponential growth rate, defined as the per capita change in the number of new cases per unit time. The reproduction number is then obtained by the formula:

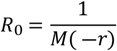

In this context, M represents the moment generating function of the generation time distribution. In a subsequent development, Obadia et *al*. [30] proposed estimating *r* through a Poisson regression of the incidences using the classical logarithm for the link function, with maximum likelihood resolution. For this application, the R library « *R0* » was developed. In this work, we will refer to this method as the *«exponential growth method»*.

#### 2.1.3. Estimation using the renewal process by Cori et al. [14]

In the absence of multiple introductions into the transmission process, Bettencourt and Ribeiro [17] used an exponential growth model to estimate the incidence of cases over a period τ :

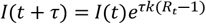

Where *1/k* represented the average infectivity period and R_t_ the effective reproduction number of time interval t.

Cori et *al*. [14] postulated that the distribution of infections was independent of generation time. Consequently, they modelled the infections produced by an infected individual at a specific point in time during a specified period using a Poisson renewal process. In this model, I(t) represents the incidence at date t, while 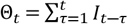 is represents the total number of cases up to and including date t. The effective reproduction number at time t, *R*_*t,τ*_ is then estimated under the assumption that the prior generation time *τ* follows a Gamma distribution with parameters a and *b*. The posterior mean and standard deviation are expressed as follows:

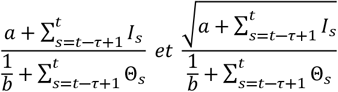

The research team developped a R library, called *« EpiEstim »*. For clarity, this method will henceforth be referred to as the *«renewal process method»*.

### 2.2. Estimation by real-time smoothing

Previous methodologies assume a complete and accurate representation of case data, which is seldom achievable in practice. Therefore, it is proposed that the reproduction rate be estimated using data smoothing techniques.

To this extent, time series of new infections for a specific disease per time unit Y_t_,t ∈ ℕ can be analyzed using a generalised additive model (GAM). The formulation for this model is presented as follows [31] :

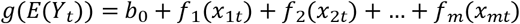

Where g represents the link function, E(Y) denotes the expected value of the dependent variable Y, b_0_ is the intercept parameter and *f*_*i*_(*x*_*it*_) are smoothing functions for the explanatory variables *x*_*it*_. In this context, these smoothing functions are defined as cubic spline functions. The cubic spline function for a variable can be expressed as follows:

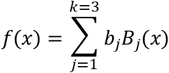

Where B_j_(x) are cubic spline basis functions and β_j_ are the coefficients to be estimated.

The use of cubic splines is justified by their piecewise continuous nature and the continuity of their first and second derivatives, which ensures the robust estimation of relationships between variables even in the presence of noise. One method to estimating the smoothing parameters is to select them in a manner that aligns 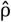 as closely as possible with the true E(Y). An appropriate measure of this closeness could be the expected mean squared error (MSE) of the model. Thus, it is reasonable to choose the smoothing parameters that minimise an estimate of this expected MSE to minimise the unbiased risk estimator (UBRE) [31,32].

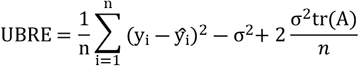

where the term n represents the number of observations, y_i_ denotes the observed value, ŷ_i_ signifies the predicted value, σ^2^ is the error variance.

The objective is to minimise the UBRE score, which requires modifying the model by adjusting the number of spline nodes. This ensures an optimal balance between model adjustment and generalization.

Once the GAM has been fitted, the smoothed function can be derived with respect to time t, enabling examination of the epidemic growth at each point.

Let F be this function.

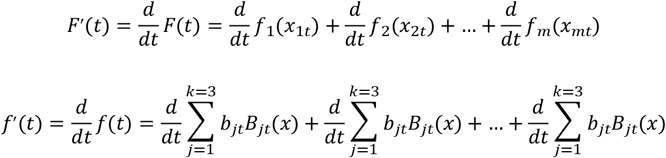

Let r represent the tangent at each time point t of the smoothing function. This tangent represents a local approximation of the epidemic’s speed at each time point. The R_t_ are related to r by the moment generating function M of the generation time distribution. The generation time distribution describes the temporal interval between the onset of symptoms in an index individual and the subsequent onset of symptoms in those they have infected.

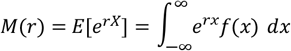

The moment generating function M(-r) of the generation time w(t) is modelled according to a gamma distribution with mean *ϱ* and standard deviation *σ* :

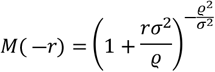

The effective reproduction number R_t_ is then defined as the inverse of this moment generating function:

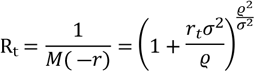

To estimate R_0_, we consider the initial phase of the epidemic up to the inflection point, which is defined as the point at which f’(t) is maximal. Subsequently, R_0_ is estimated as the median of the values of R_t_ during the initial phase.

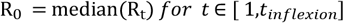

Where t_inflexion_ is the time corresponding to the inflection point, at the epidemic onset.

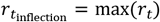

To estimate R_0_, for a given scenario, the median of the medians of the R_t_ for each simulation is calculated, with the 2.5% and 97.5% quantiles of the associated estimates serving as the bounds of the tolerance interval at the 95% level (TI_95%_). In this work, we will refer to this method as the *« splines smoothing method»*.

The supplementary file (S4 File) document provides a comprehensive illustration of this method.

### 2.3. Data and simulation

#### 2.3.1. Simulation scenario

The data presented here were derived from an epidemiological simulation, which illustrates the spread of measles. The duration of the epidemic varied between one and two years, depending on the scenario under consideration. Three distinct scenarios were established. To ensure a precision of 0.01 and a variance of 0.09, in accordance with best practices in simulation [33,34], a total of 3458 simulations were performed for each scenario. Each simulation was conducted on a population of 20 million individuals.

The infection process was modelled using a SEIR method, as illustrated in figure 1 (Fig 1). Considering the clinical knowledge available regarding measles, the transmission dynamics can be described by the compartmental model presented below:

**Figure 1:**
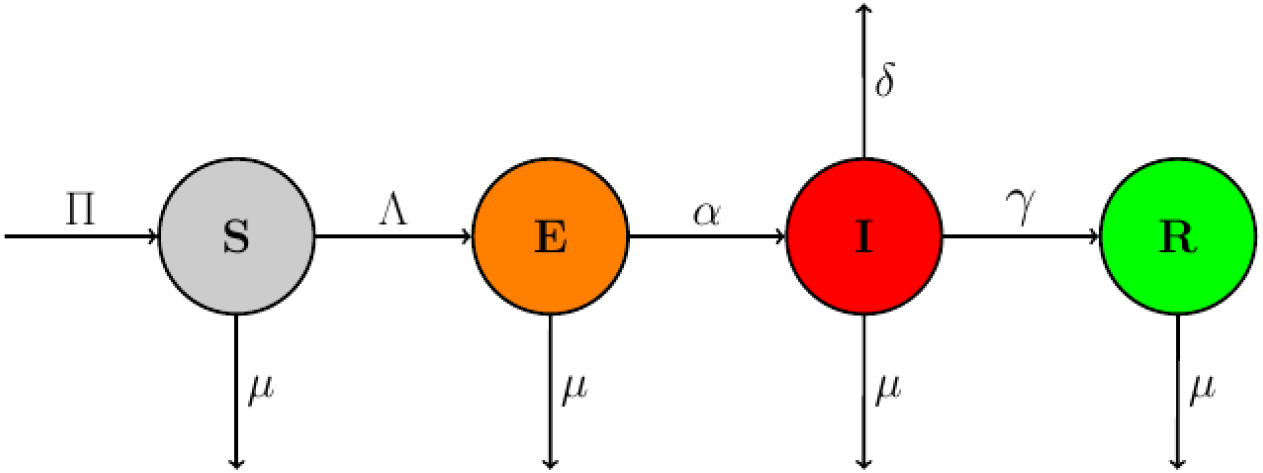
Transmission model. The figure represents a schematic of the SEIR model, in which S represents susceptible individuals, E represents exposed (infected but not infectious) individuals, I represents infectious individuals (who can spread the pathogen), and R represents Recovered individuals (who cannot be infected anymore). The arrows indicate the transitions between the population states, with the corresponding rates. A The inverse of incubation time; γ the inverse of infectivity time; µ natural mortality rate; δ measles mortality rate; Π recruitment rate; birth rate; Λ no infectious infected people.

The SEIR compartmental model was transformed by the addition of a stochastic component to the system of ordinary differential equations, with the objective of simulating several given datasets on the measles epidemic. The stochastic simulation model, which was the subject of our subsequent analysis, was as follows:

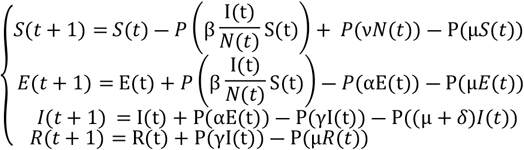

The specific parameters are outlined in Table 1.

**Table I:** The parameters of the transmission model.

| Parameters | Description |
| --- | --- |
| S | Susceptible |
| E | Exposed |
| I | Infected |
| R | Recovered |
| $N = S + E + I + R$ | Total population |
| $\alpha$ | Incubation time |
| $\gamma$ | Infectivity time |
| $\mu$ | Natural mortality rate |
| $\nu$ | Birth rate |
| c | The rate of encounters in the population |
| p | Probability of contamination following an encounter with an infected person |
| $\delta$ | Disease mortality rate |
| $\Pi = \nu * N$ | Recruitment rate |
| $\beta = cp$ | The transmissibility of the disease |
| $\Lambda = \beta \frac{S}{N} I$ | Non-infectious infected people |

In this context, the variable P represented a standard Poisson processing procedure. The supplementary file (S2 File) describes the simulation algorithm.

During the simulations, the number of new infected cases per day was recorded for each scenario. The number of knots of the spline were measured on a single data set per scenario, randomly selected, were extrapolated to the others after a sensitivity analysis.

#### 2.3.2. Evaluation criteria

To assess and compare estimates and methods, four performance indicators were used, as recommended [33,34]. These indicators are measured across all the provided datasets within each scenario. The supplementary file (S3 File) offers a detailed description of the characteristics and specifications of each indicator: the estimated bias (Bias(R_0_)), the mean square error (MSE), the coverage rate (CR), the significance rate (SR) and the false negative rate (FNR).

#### 2.3.3. Transmission model parameters

The parameters used in this model are listed in Table 2, which includes their descriptions and sources.

**Table II:** Transmission model parameters.

| Parameters | Values | Description |
| --- | --- | --- |
| $R_0$ | 11.52; 2.88; 1.06 | A uniform random draw from 10.4-12 with two values at the end of the epidemic. In a systematic review on measles $R_0$, was 10.4 in low birth-rate and 12.9 in high birth-rate [35]. |
| $\alpha$ | 10 | The incubation period for measles is typically 10 days [36,37]. |
| $\gamma$ | 8 | The contagious period is defined as starting four days before the onset of the rash (14 days) and continuing until four days afterward [36,37]. |
| N | 20 millions | A total population of 20 million is considered. |
| $\mu$ | 9.7 | The natural mortality rate is 9.7 per 1000 people. |
| $\nu$ | 10.1 | The birth rate is 10.1 per 1000 people. |
| $\delta$ | 4.2 | Some individuals may die from complications associated with measles, though such cases are rare and unlikely [36,37]. The measles mortality rate is estimated to be 4.2% [38]. |
| T | 365 jours | We will simulate an epidemic lasting one year (365 days). |

The supplementary document (S1 File) contains a detailed account of the determination of the basic reproduction number considering the compartmental model based on ordinary differential equations. The expression of *R*_0_ is determined as follows:

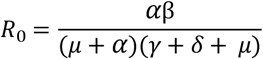

Since the only unknown variable in this equation is β, we adjust its value to achieve the desired R_0_ for our different scenarios.

In a systematic review on measles R_0_, stratifying by birth rate, median measles R_0_ was 10.4 in low birth-rate settings and 12.9 in high birth-rate settings [35]. The study’s R_0_ values was obtained by a uniform random draw from the interval with two values at the end of the epidemic.

## 3. Results

### 3.1. Data

The change in the number of new infected cases over time (in days) is shown for each scenario.

A total of 10,000 one-year time series were generated for each scenario. After excluding simulations that did not exhibit sufficient epidemic dynamics (defined as an epidemic duration of less than 15 days or data with constant values throughout the epidemic period), the remaining 8,900 series were retained for further analyses.

Figure 2, panel a (scenario 1, R_0_ =11.52 illustrates a sharp increase in the number of cases, peaking around day 100 with approximately 6 million cases. Following this peak, the number of cases declines steeply, reaching a low point around day 200 (Fig 2). This distribution indicates an epidemic characterized by a rapid initial spread followed by an equally swift resolution.

**Figure 2:**
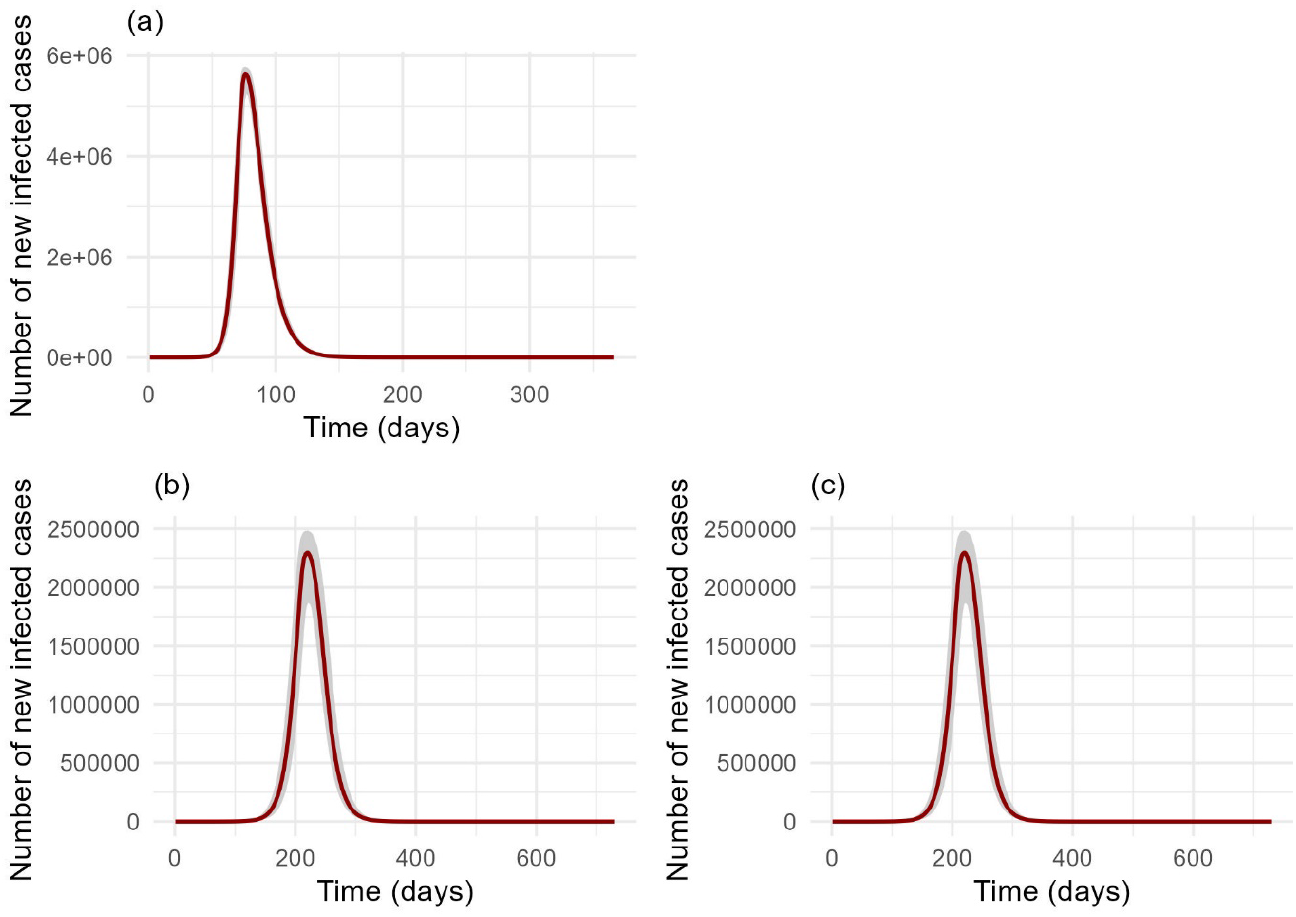
Evolution of the median daily infected cases from the simulation data, with the inter-quartile ranges included. The change in the number of new infected cases over time (in days) is shown for each scenario. Each value corresponds to the median number of cases per day calculated based on the simulations carried out in the scenario in question. This makes it possible to present the dynamics of the epidemic in a synthetic way, considering the fluctuations observed in the different simulations, while at the same time providing an overview of the main trends. The number of cases is represented on the x-axis, and the time in days is represented on the y-axis.

Figure 2, panel b (scenario 2, R_0_ =2.88) depicts a distribution similar to scenario 1, but with a delayed and lower peak of approximately 2.5 million cases around day 200, followed by a return to near-zero levels around day 300 (Fig 2).

Figure 2, panel c (scenario 3, R_0_ =1.06) depicts a more irregular curve, exhibiting multiple minor peaks throughout the 365-day period. The number of cases fluctuates between 1 and 4 suggesting that the epidemic may have been characterised by sporadic transmission or successive waves of low-intensity transmission (Fig 2).

### 3.2. Estimations of R_0_

#### Scenario 1, R_0_ = 11.52

The figure illustrates the estimated value of R_0_, as determined by the three different methods used in scenario 1. For the renewal process method, the median estimate of R_0_ is approximately 12.78, with a tolerance interval of 10.48 to 17.48 (Table 3, Fig 3). This estimate has a bias of 1.69, with an MSE of 5.65. The splines smoothing method gives a median estimate of R_0_ of approximately 12.33, with a tolerance interval from 8.34 to 18.03, a bias of 1.19 and an MSE of 7.31(Table 3, Fig 3). The exponential growth method provides a median estimate of R_0_ for of approximately 13.90, with a tolerance interval from 10.39 to 33.47, a bias of 4.17 and an MSE of 47.41 (Table 3, Fig 3). For the coverage rate (CR), the renewal process method achieved 75.55%, the splines smoothing method 97.04%, and the exponential growth method 56.29% (Table 3). All estimates from these methods were statistically significant (SR = 100) with a false negative rate (FNR) of zero (Table 3).

**Table III:**
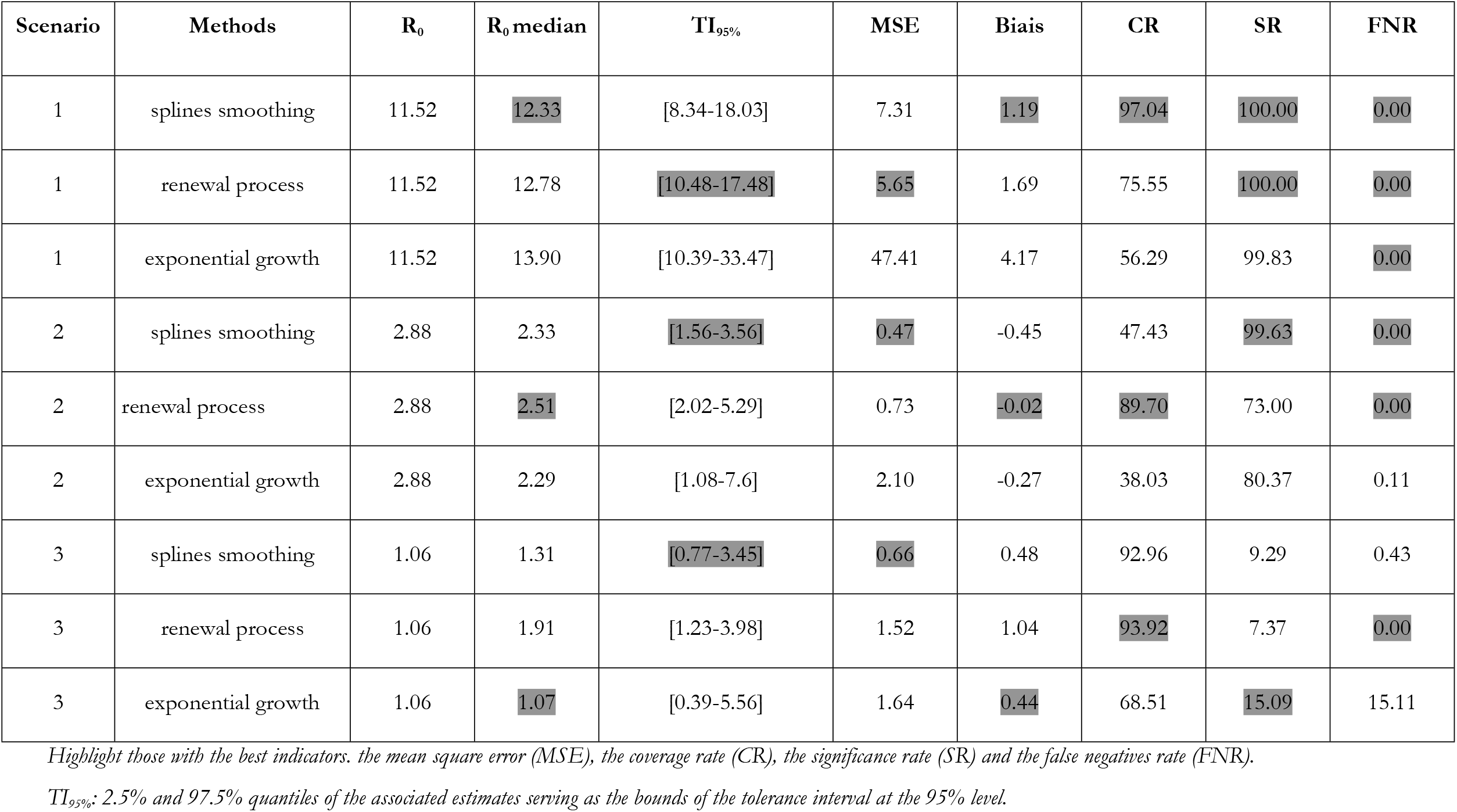
Performance indicators for estimating R_0_.

**Figure 3:**
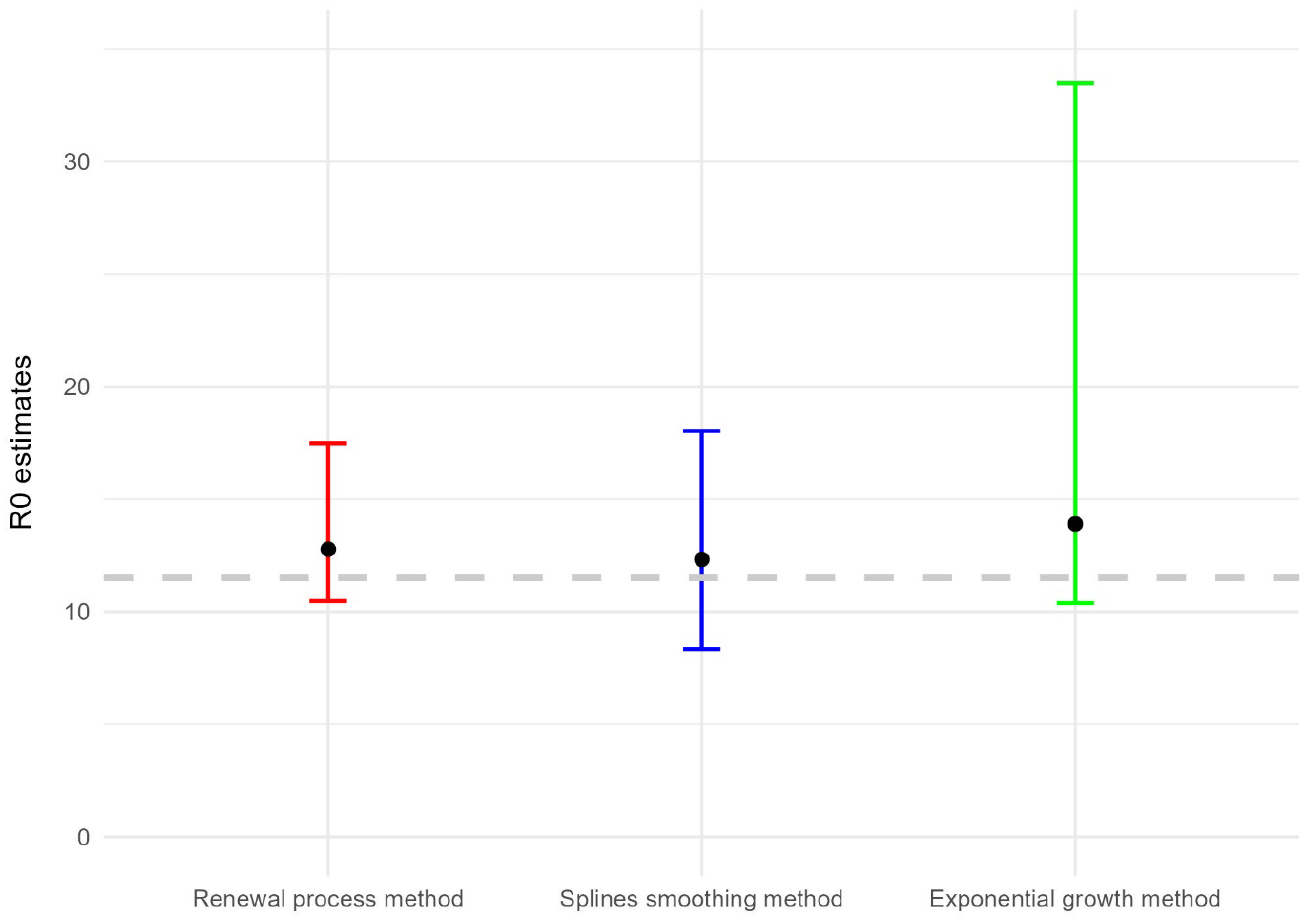
Estimation of R_0_ with its associated tolerance interval in scenario 1 (R_0_=11.52). The figure depicts the estimated value of R_0_, along with its associated tolerance interval, obtained through the application of three distinct methodologies. Each data point represents the median value of R_0_, calculated from the respective simulations, while the upper and lower bounds represent the quantiles at 2.5% and 97.5% of the estimated values, respectively. The grey horizontal line denotes the actual value of R_0_ utilized in the simulations.

Figure 4 illustrates the distribution of R_0_ estimates derived from the three distinct methods in a scenario where R_0_ is set to 11.52. The three estimation methods (splines smoothing method, renewal process method and exponential growth method) produce distributions of R_0_ estimates with comparable shapes, exhibiting a slight asymmetry towards the right. The exponential growth method appears to yield slightly higher estimates on average (15.69) than the other two methods, with values of 13.20(renewal process), and 12.70 (splines smoothing). Despite these differences, the dispersion of the estimates is similar across all three methods. These findings suggest that while the methods perform comparably in capturing underlying trends in the data, minor discrepancies in average estimated may occur (Fig 4).

**Figure 4:**
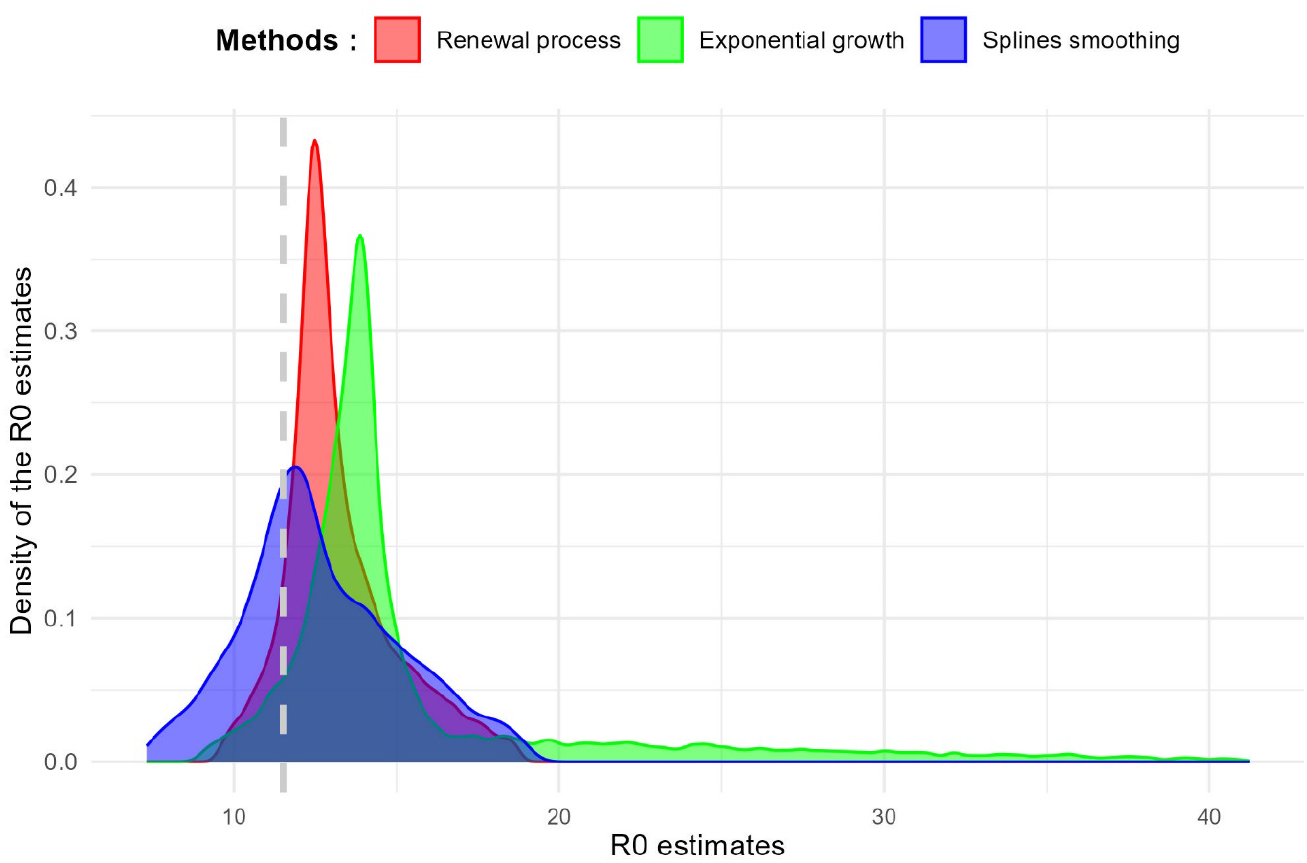
Distribution of the R_0_ estimate in scenario 1 (R_0_=11.52). The figure illustrates the distribution of R_0_ estimates derived from the three methods. The horizontal axis represents the estimated R_0_ values, while the vertical axis corresponds to the density of the estimates obtained. The vertical grey line denotes the actual value of R_0_ employed in the simulations.

#### Scenario 2, R_0_= 2.88

Figure 5 illustrates the R_0_ estimates derived from the three methods for scenario 2. The renewal process method yielded a median estimate of R_0_ of 2.51, with a tolerance interval from 2.02 to 5.29 (Table 3, Fig 5). The splines smoothing method yielded a median estimate of R_0_ of 2.33, with a tolerance interval of 1.56 to 3.56 (Table 3, Fig 5). For the exponential growth method, the median estimate R_0_ was 2.29 with a tolerance interval from 1.08 to 7.6 (Table 3, Fig 5). The observed biases for these estimates were −0.02 for the renewal process method, −0.45 for the splines smoothing and −0.27 for the exponential growth method, with MSE of 0.7, 0.47 and 2.10, respectively (Table 3). In terms of CR, the renewal process method achieved 89.70%, the splines smoothing method 47.43%, and the exponential growth method 38.03%. The SR were 73.00% for the renewal process method, 99.63% for splines smoothing and 80.37% for the exponential growth method (Table 3). The FNR was zero for both the renewal process method and splines smoothing, while it was 0.11% for the exponential growth method.

**Figure 5:**
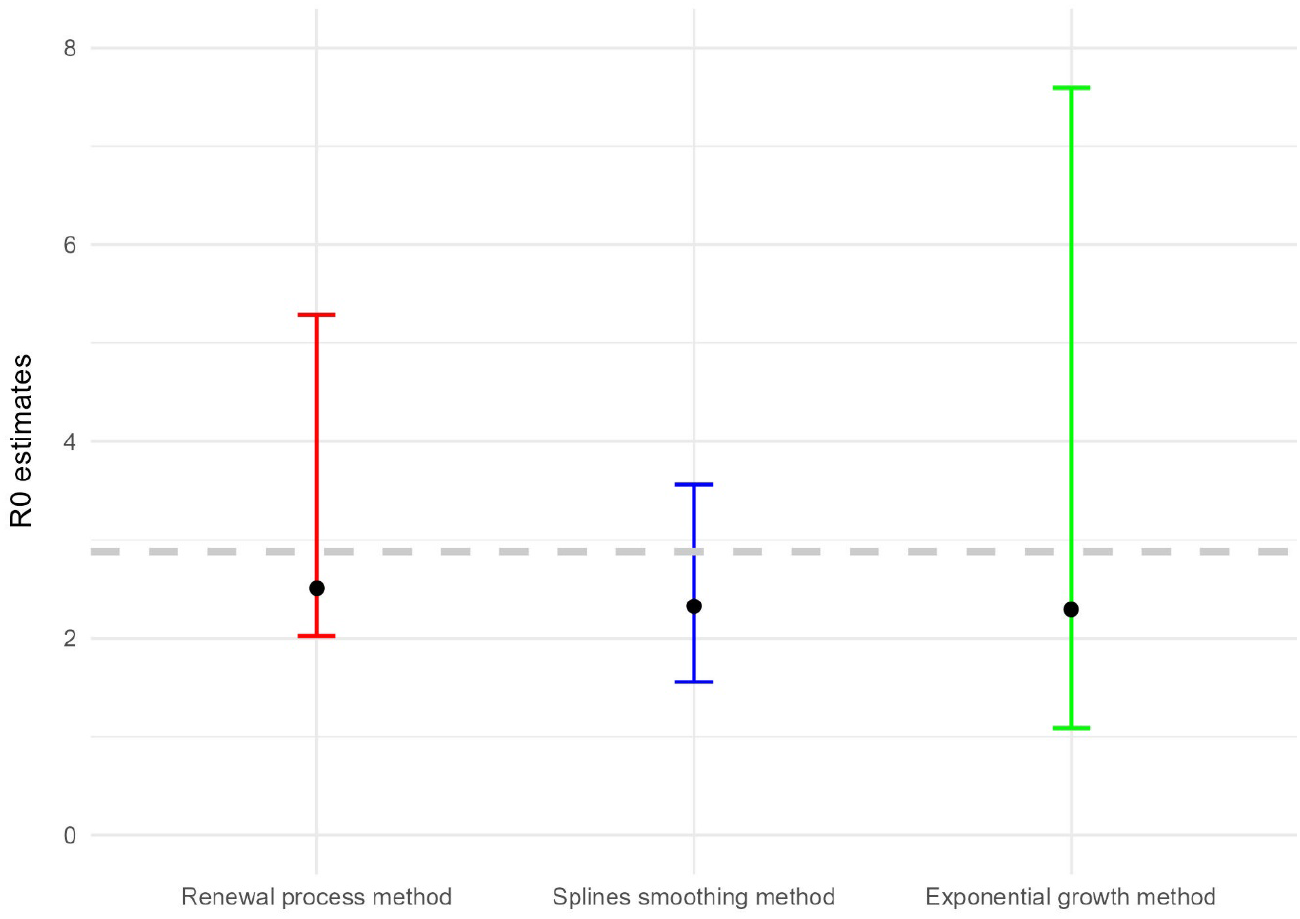
Estimation of R_0_ with its associated tolerance interval in scenario 2 (R_0_=2.88). The figure depicts the estimated value of R_0_, along with its associated tolerance interval, obtained through the application of three distinct methodologies. Each data point represents the median value of R_0_, calculated from the respective simulations, while the upper and lower bounds represent the quantiles at 2.5% and 97.5% of the estimated values, respectively. The grey horizontal line denotes the actual value of R_0_ utilized in the simulations.

Figure 6 illustrates the distribution of R_0_ estimates obtained using the three methods applied to scenario 2. There are slight differences in uncertainties and biases across the methods. The point estimate values exhibit slight variability across the three methods with the splines smoothing method producing distributions with more concentration around the true value. Both the renewal process method and the exponential growth method display a rightward skew, with the exponential growth method showing an even more pronounced tail than the renewal process method. Overall, it can be observed that all estimates are slightly below the true value (Fig 6).

**Figure 6:**
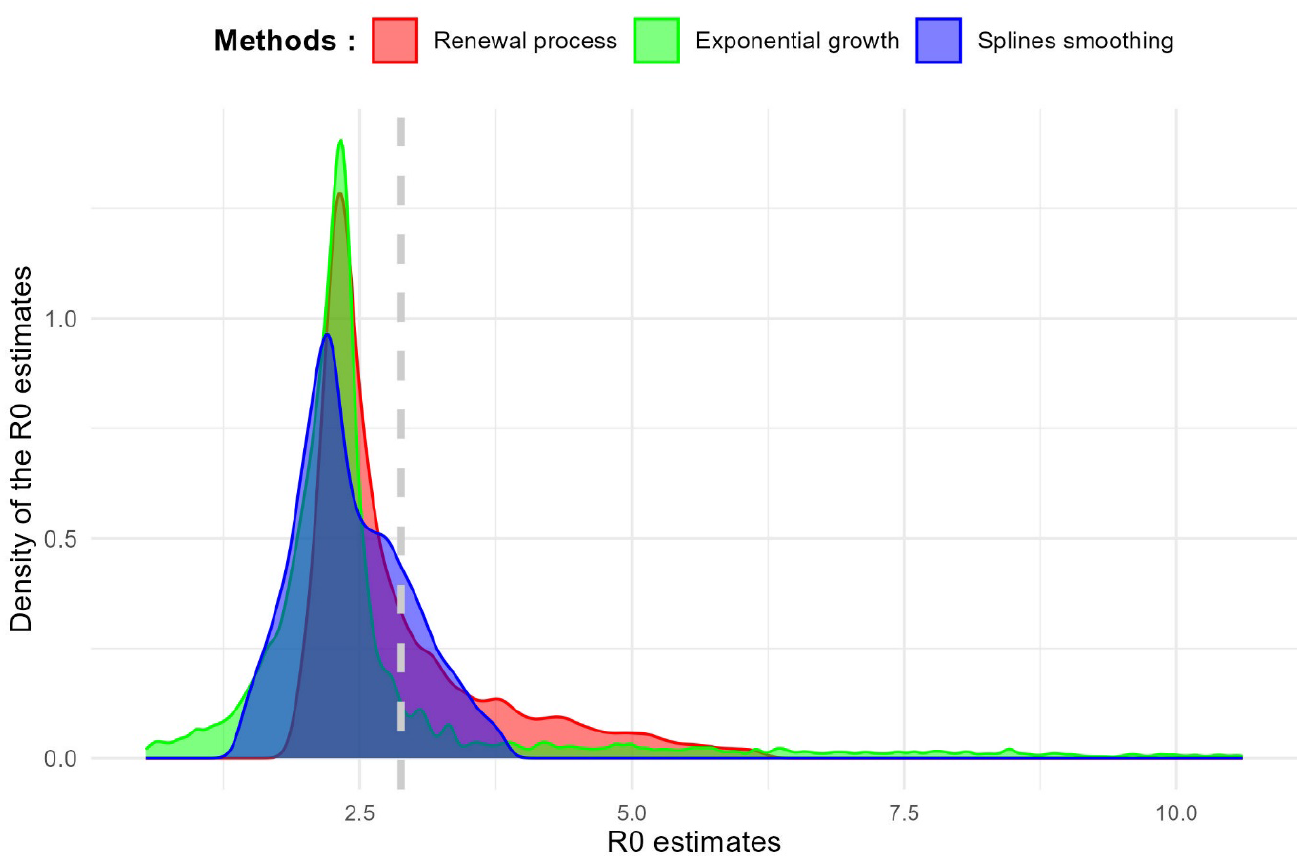
Distribution of the R_0_ estimate in scenario 2 (R_0_=2.88). The figure illustrates the distribution of R_0_ estimates derived from the three methods. The horizontal axis represents the estimated R_0_ values, while the vertical axis corresponds to the density of the estimates obtained. The vertical grey line denotes the actual value of R_0_ employed in the simulations.

#### Scenario 3, R_0_ = 1,06

Figure 7 illustrates the estimated value of R_0_ derived from the various methods applied in scenario 3, where the actual value of R_0_ is 1.06 (illustrated by the grey dotted line).

**Figure 7:**
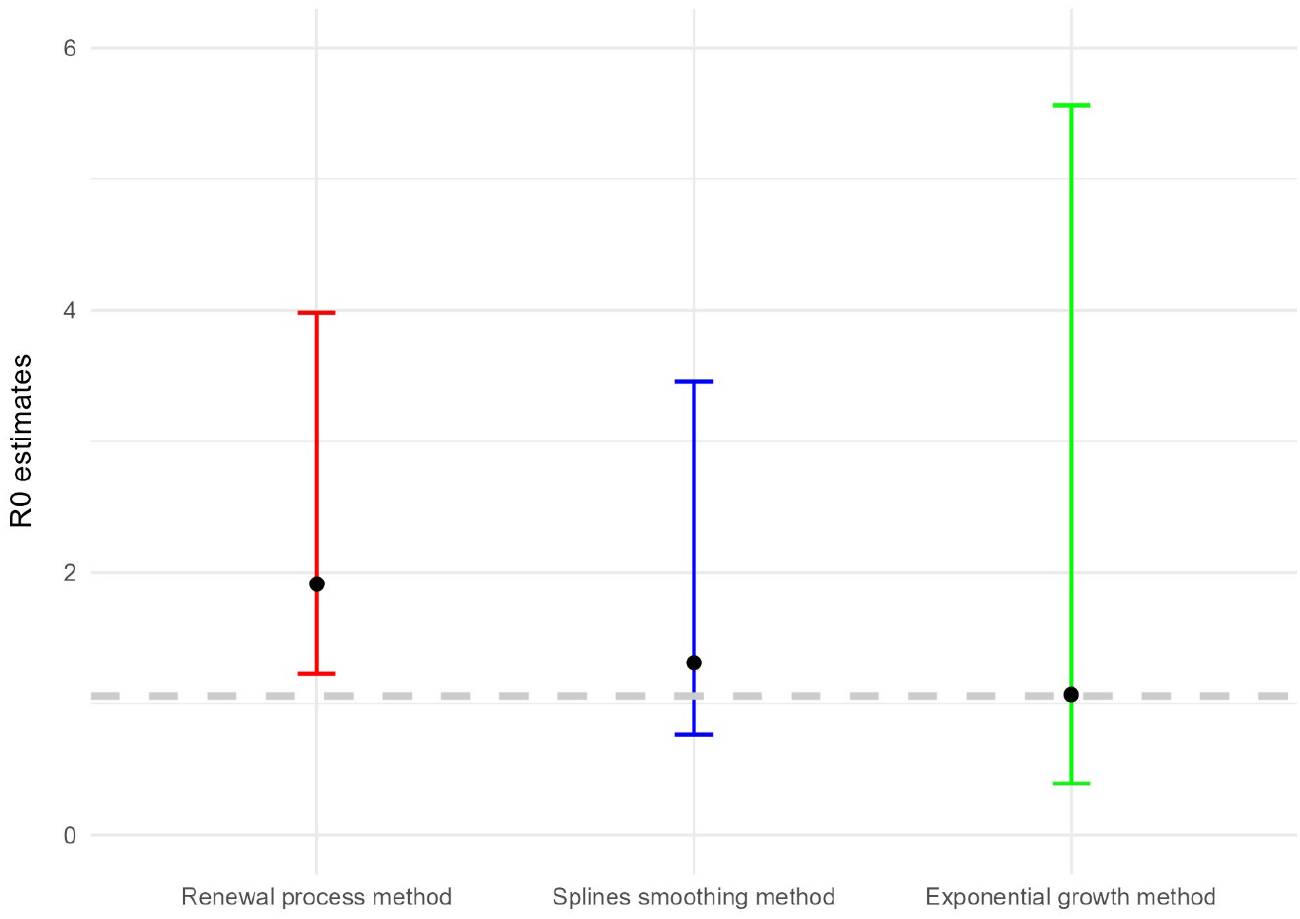
Estimation of R_0_ with its associated tolerance interval in scenario 3 (R_0_=1.06). The figure depicts the estimated value of R_0_, along with its associated tolerance interval, obtained through the application of three distinct methodologies. Each data point represents the median value of R_0_, calculated from the respective simulations, while the upper and lower bounds represent the quantiles at 2.5% and 97.5% of the estimated values, respectively. The grey horizontal line denotes the actual value of R_0_ utilized in the simulations.

The renewal process method yielded a median estimate of R_0_ of 1.91 with a tolerance interval of 1.23 to 3.98 (Table 3, Fig 7). The observed bias is 1.04, and the MSE is 1.52. For the CR, the renewal process method achieved 93.92%, with a SR of 7.37% and a FNR of 0% (Table 3). The splines smoothing method yielded a median R_0_ estimate of 1.31 with a tolerance interval from 0.77 to 3.19 (Table 3, Fig 7). The observed bias is 0.48, and the MSE is 0.66 (Table 3). The CR is 92.96%, the SR is 9.29%, and the FNR is 0.43%. The exponential growth method yielded an estimated R_0_ at 1.64 with a tolerance interval of 0.39 to 5.56 (Table 3, Fig 7). The observed bias is 0.44, and the MSE is 1.64. The CR was 68.51%, and the SR was 15.09%, with a FNR of 15.11% (Table 3).

Figure 8 illustrates the distribution of R_0_ estimates obtained from the three methods used in scenario 3, all of which display a general bell-shaped pattern. The splines’ smoothing method has concentration around the true value, suggesting relatively consistent estimates. The renewal process method appears to yield slightly higher estimates of R_0_ than the splines smoothing method. Meanwhile, the exponential growth method’s distribution is somewhat broader than the other two methods (Fig 8).

**Figure 8:**
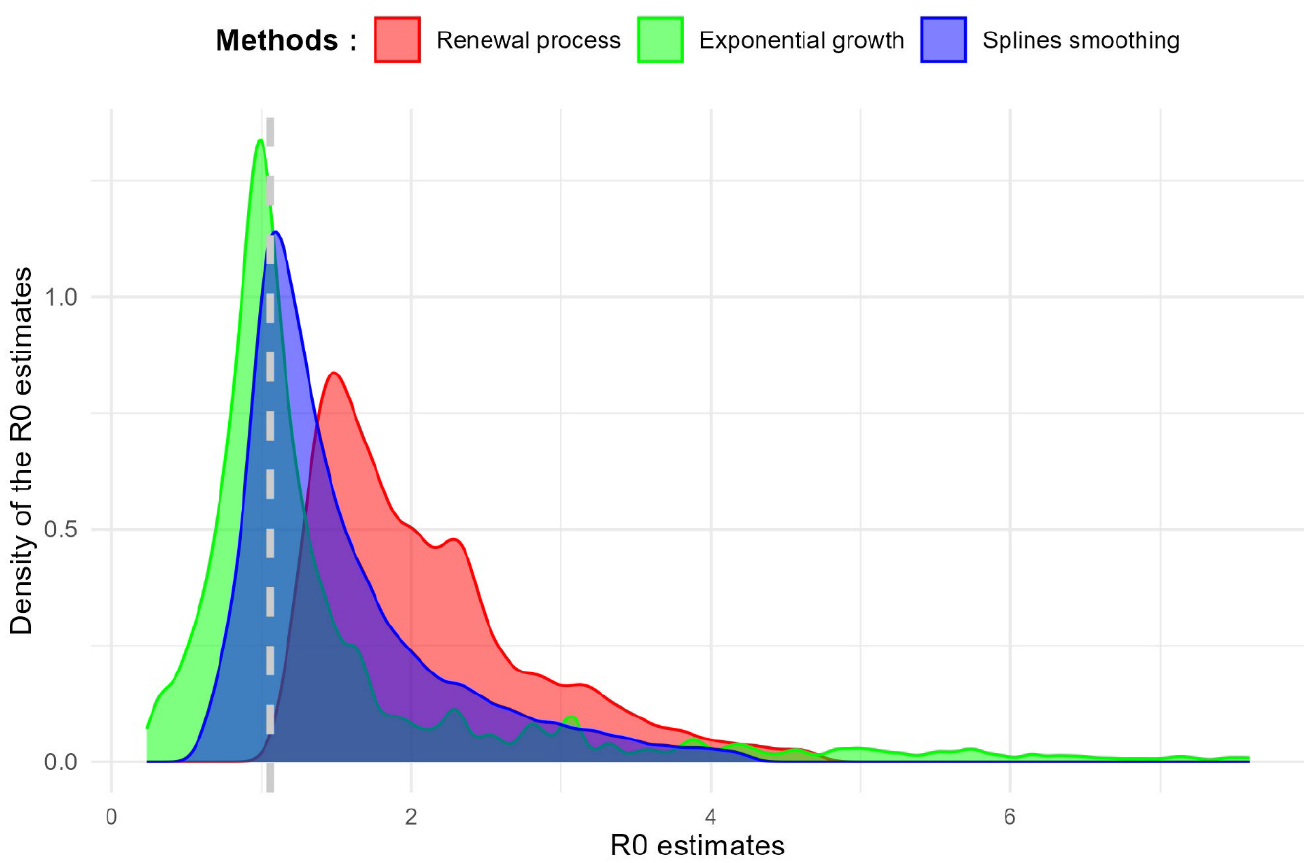
Distribution of the R_0_ estimate in scenario 3 (R_0_=1.06). The figure illustrates the distribution of R_0_ estimates derived from the three methods. The horizontal axis represents the estimated R_0_ values, while the vertical axis corresponds to the density of the estimates obtained. The vertical grey line denotes the actual value of R_0_ employed in the simulations.

### 3.3. Model performance measures for R0

Table 3 summarizes the performance of the different methods for R_0_ estimation across the three scenarios. The renewal process method, the splines smoothing method, and the exponential growth method yielded varying estimates of R_0_ highlighting distinct performance characteristics across the scenarios.

The R_0_ estimates from the renewal process method are relatively close to the true value, with narrower tolerance intervals. This method demonstrates a moderate MSE and a slightly elevated bias. The CR is generally high. This method has a zero FNR.

The splines smoothing method has wider tolerance intervals than those from the renewal process method. It generally has lower MSE and bias. The FNR for this method is zero in all scenarios except in scenario 3 (0.43%, Table 3).

The exponential growth method produces estimates with considerable variability. The mean square error is higher compared to the other methods. Additionally, the bias is moderately elevated. The CR is lower. Furthermore, the FNR is non-zero in scenarios 2 and 3, reaching 15.11% (Table 3).

### 3.4. Estimation of R_t_ by splines smoothing and renewal process methods

The change in the number of new infected cases over time (in days) is shown for each scenario, with the median of the different quartiles of the epidemic duration corresponding to the three time points used for R_t_ analysis (Fig 9 in appendix).

**Figure 9:**
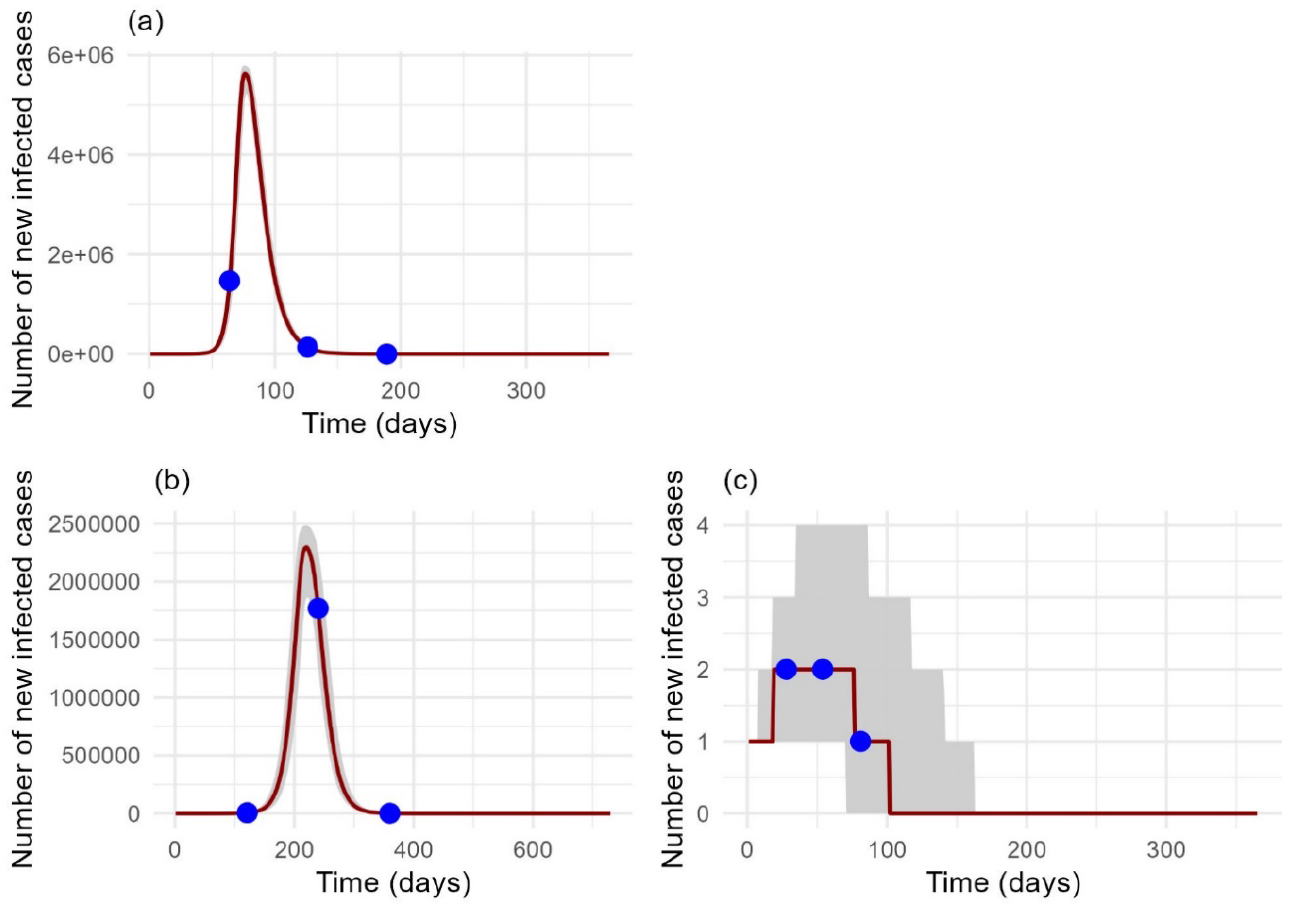
Evolution of the median daily infected cases from the simulation data, with the inter-quartile ranges included. The change in the number of new infected cases over time (in days) is shown for each scenario. Each value corresponds to the median number of cases per day calculated based on the simulations carried out in the scenario in question. The number of cases is represented on the x-axis, and the time in days is represented on the y-axis. The different points correspond to the median of the quartiles of the epidemic duration in each simulation.

In general, the two methods demonstrate comparable performance. However, for the first quartile of scenario 1, corresponding to the ascending phase of the epidemic, values below 1 were observed for the renewal process method (Table 4).

**Table IV:** Performance indicators for estimating R_t_.

| Scenario | Quartile | Methods | $R_0$ | $R_t$ median | $TI_{95\%}$ | SR | FNR |
| --- | --- | --- | --- | --- | --- | --- | --- |
| 1 | Q1 | splines smoothing | 11.52 | 9.14 | [1.73-12.44] | 99.2 | 0.8 |
| 1 | Q1 | renewal process | 11.52 | 4.28 | [0.92-11.22] | 96.78 | 3.21 |
| 1 | Q2 | splines smoothing | 11.52 | 0.31 | [0.3-0.32] | 0 | 100 |
| 1 | Q2 | renewal process | 11.52 | 0.30 | [0.29-0.31] | 0 | 100 |
| 1 | Q3 | splines smoothing | 11.52 | 0.31 | [0.24-0.41] | 0 | 99.85 |
| 1 | Q3 | renewal process | 11.52 | 0.25 | [0.13-0.31] | 0.15 | 98.89 |
| 2 | Q1 | splines smoothing | 2.88 | 2.34 | [2.25-2.42] | 99.58 | 0.04 |
| 2 | Q1 | renewal process | 2.88 | 2.30 | [2.14-2.36] | 99.85 | 0 |
| 2 | Q2 | splines smoothing | 2.88 | 0.69 | [0.5-1.30] | 9.19 | 90.40 |
| 2 | Q2 | renewal process | 2.88 | 0.64 | [0.49-1.14] | 5.16 | 94.64 |
| 2 | Q3 | splines smoothing | 2.88 | 0.47 | [0.43-0.51] | 0.01 | 99.57 |
| 2 | Q3 | renewal process | 2.88 | 0.44 | [0.34-0.47] | 0.44 | 99.24 |
| 3 | Q1 | splines smoothing | 1.06 | 1.21 | [0.25-3.95] | 17.78 | 3.37 |
| 3 | Q1 | renewal process | 1.06 | 0.36 | [0.01-1.61] | 69.87 | 0.71 |
| 3 | Q2 | splines smoothing | 1.06 | 1.01 | [0.2-3.77] | 8.37 | 8.57 |
| 3 | Q2 | renewal process | 1.06 | 0.32 | [0.01-1.39] | 60.13 | 1.75 |
| 3 | Q3 | splines smoothing | 1.06 | 0.87 | [0.18-4.03] | 4.78 | 16.85 |
| 3 | Q3 | renewal process | 1.06 | 0.22 | [0.01-1.19] | 53.35 | 2.33 |

SR: the significance rate, FNR : the false negatives rate, $Q1$ : median of first quartiles, $Q2$ : median of second quartiles, $Q3$ : median of third quartiles. The different quartiles correspond to the quartiles of the epidemic duration in each simulation. $TI_{95\%}$ : 2.5% and 97.5% quantiles of the associated estimates serving as the bounds of the tolerance interval at the 95% level.

## 4. Discussion

The three methods yielded varying estimates of R_0_ each accompanied by different tolerance intervals and distributions across the three scenarios. Each method demonstrated unique strengths and limitations regarding precision, variability, bias, and the ability to detect the presence of an epidemic. These findings underscore the importance of selecting an appropriate method based on the specific epidemiological context and the attributes of the available data.

The renewal process method offered relatively accurate R_0_ estimates, with tighter tolerance intervals. However, this was accompanied by a slight increase in bias and a moderate MSE, which may indicate a risk of systematic deviation and underestimation of extremes in epidemic dynamics. The exponential growth method generates estimates with considerable variability and a higher MSE, indicating greater uncertainty in the estimates. The bias is moderately high, which may indicate a tendency to overestimate R_0_ in the estimates. The CR is lower, suggesting that the tolerance intervals are less frequently aligned with the true value of R_0_. Moreover, this method is unable to correctly identify the presence of an epidemic, particularly in cases where the R_0_ is low. The splines smoothing method struck an effective balance between bias, MSE and CR, demonstrating its capability to capture epidemic dynamics. It generally exhibited low MSE and bias, providing good precision in its estimates. By utilizing splines to model non-linear relationships between variables, this method can accommodate intricate fluctuations in the data set [31].

Both the splines smoothing method and the renewal process method demonstrate comparable levels of uncertainty and efficacy in epidemic detection, with minor discrepancies in their respective biases and false-negative rates when estimating R_t_. It is evident that neither method exhibits a distinct superiority across all contexts. Their efficacy varies depending on performance priorities. The choice of method may depend on the emphasized criteria, such as minimizing the MSE or reducing the FNR, particularly in scenarios where R_0_ is low, like scenario 2 and 3. The efficacy of the renewal process method, developed by Cori et *al*. [14,39], has been demonstrated by Gostic et *al*. in comparison with other methods [40]. Similarly, the exponential growth method, developed by Obadia et *al*., has also demonstrated its efficacy in detecting epidemics [30].

Furthermore, the spline smoothing method has demonstrated an ability to identify the initial phase of an epidemic more effectively. The initial phase of an epidemic crucial for estimating R_0_, regardless of the method used [14,30,39,40]. This period is characterised by transmission occurring in a manner that is most akin to its natural dynamics, prior to the implementation of interventions that have the potential to significantly impact the spread of the disease. In the event of an inadequate identification of this phase, the resulting estimates of R_0_ and R_t_ may be subject to bias or imprecision. The renewal process method identifies the initial phase implicitly by selecting the time window over which the first incidence data are analysed. It is typically incumbent upon the user to specify this period. The renewal process method permits the calculation of R_t_ over a series of successive time windows. Consequently, the user can modify the specified time window to encompass the initial phase of exponential growth. To ascertain the exponential growth method, it is necessary for the user to manually select the portion of the epidemic curve that represents the initial phase. This is typically accomplished by examining the exponential component of the incidence curve. In contrast to the renewal process method and the exponential growth method which frequently necessitate explicit or manual identification of the initial phase, the splines smoothing method offers a greater flexibility in modelling the initial phase without imposing constraints on the duration or characteristics of this phase. The smoothing properties of the splines smoothing method enable flexible identification of the inflection point of the epidemic, which may be used as the end of the period of exponential growth. In this context, the phase is defined up to the inflection point, where f’(t) reaches its maximum.

Furthermore, as it is founded upon the principles of smoothing, the splines smoothing method affords the flexibility to utilise disparate smoothing techniques contingent upon the attributes of the data, thus facilitating a comprehension of the epidemic’s dynamics. In contrast to the renewal process method and the exponential growth method, the splines smoothing method can adopt a range of distributions, including the Poisson distribution, the negative binomial distribution, and the Gaussian distribution, in accordance with the specific characteristics of the data and the epidemiological context [31,41]. Furthermore, the splines smoothing method could also incorporate covariates to adjust estimates [31,42,43]. For instance, covariates such as public health interventions, seasonal variations, and other contextual factors can be integrated into the model, thereby facilitating a more comprehensive analysis that is tailored to the specific circumstances of the epidemic.

## 5. Conclusion

The study introduces an effective method for calculating R_0_ using a smoothing technique. The findings have significant implications for public health and methodology, particularly concerning surveillance, intervention, and epidemic management. The spline smoothing method can adapt to data from various contexts with differing epidemic dynamics, which is essential for effective surveillance in environments with fluctuating transmission rates. The precise yet straightforward identification of the initial phase by the spline smoothing method supports proactive adjustments in control and prevention measures, representing a noteworthy contribution to the study of epidemic dynamics. The ability to use different smoothing techniques based on the data’s characteristics allows for a deeper understanding of epidemic dynamics.

## Data Availability

All data and statistical analysis code are available on Zenodo at link : 10.5281/zenodo.14645488. Sample R code is provided for the purpose of enabling users to reproduce the results or apply the method to their own data. The code includes the principal steps for fitting the transmission model and estimating the parameters with the proposed methods. All data and statistical analysis code are available on Zenodo at link : 10.5281/zenodo.1464599.

https://doi.org/10.5281/zenodo.14645488

https://doi.org/10.5281/zenodo.1464599

## Data availability statement

All data and statistical analysis code are available on Zenodo at link : XXXXXX.

## Contributors

JG developed the original idea of the study.

MB, JG, SMAS, and CSB designed the study.

MB conducted the simulations and analyses under the supervision of JG and SMAS.

BR and CSB contributed to the development of the simulation code.

BEAD contributed to the mathematical development of the epidemiological system used for the simulation.

BK contributed to the drafting of the manuscript and interpretation of the data.

AFS provided the necessary epidemiological expertise for the study and interpreted the data.

MB wrote the first version of the manuscript.

All authors contributed to the revision and editing of the manuscript.

## Acknowledgments

We would like to express our deep gratitude to Cédric Pennetier for his scientific, administrative, and personal support throughout this work.

We would like to express our deep gratitude to the “Institut de Recherche en Sciences de la Santé (IRSS)”, the REACT2 project and the “Institut de Recherche pour le Développement (IRD)”, Representation in Burkina Faso for awarding us the thesis scholarship, that enabled us to conduct this research. We are also indebted to them for their unwavering support throughout this endeavor.

Additionally, I would like to express my gratitude to the French Embassy in Burkina Faso for providing me with a mobility grant, which greatly facilitated my research.

The financial and scientific support provided by these institutions was instrumental in the success of this project, and we are sincerely grateful for this opportunity.

## Supporting information

**Fig. S1: Evolution of the median daily infected cases from the simulation data, with the inter-quartile ranges included**

**<u>S1 File:</u> A mathematical analysis of the transmission model**: This section offers a comprehensive account of the mathematical model employed for the analysis of disease transmission.

**<u>S2 File:</u> Model algorithm for generating simulation data**: This section outlines the algorithmic method employed for the generation of synthetic data, which is subsequently employed for the assessment and validation of estimation methodologies.

**<u>S3 File:</u> Definition of judgement criteria:** This section provides a comprehensive account of the criteria employed to assess the efficacy of the model and estimation techniques, including the estimated bias 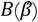, the mean square error (MSE), the coverage rate (CR), the significance rate (SR) and the false negatives (FNR).

**<u>S4 File:</u> Method implementation**: Sample R code is provided for the purpose of enabling users to reproduce the results or apply the method to their own data. The code includes the principal steps for fitting the transmission model and estimating the parameters with the proposed method.

## References

1. Brizzi A, O’Driscoll M, Dorigatti I. Refining Reproduction Number Estimates to Account for Unobserved Generations of Infection in Emerging Epidemics. Clin Infect Dis Off Publ Infect Dis Soc Am. 17 févr 2022;75(1):e114–21.

2. Somda SMA, Ouedraogo B, Pare CB, Kouanda S. Estimation of the Serial Interval and the Effective Reproductive Number of COVID-19 Outbreak Using Contact Data in Burkina Faso, a Sub-Saharan African Country. Comput Math Methods Med. 2022;2022:8239915.

3. Koutou O, Diabaté AB, Sangaré B. Mathematical analysis of the impact of the media coverage in mitigating the outbreak of COVID-19. Math Comput Simul. mars 2023;205:600–18.

4. Iyaniwura SA, Rabiu M, David JF, Kong JD. The basic reproduction number of COVID-19 across Africa. PloS One. 2022;17(2):e0264455.

5. Liu Y, Tang JW, Lam TTY. Transmission dynamics of the COVID-19 epidemic in England. Int J Infect Dis IJID Off Publ Int Soc Infect Dis. mars 2021;104:132–8.

6. Mallela A, Neumann J, Miller EF, Chen Y, Posner RG, Lin YT, et al. Bayesian Inference of State-Level COVID-19 Basic Reproduction Numbers across the United States. Viruses. 15 janv 2022;14(1):157.

7. Fraser C. Estimating Individual and Household Reproduction Numbers in an Emerging Epidemic. PLOS ONE. 22 août 2007;2(8):e758.

8. Cintrón-Arias A, Castillo-Chávez C, Bettencourt LMA, Lloyd AL, Banks HT, Cintrón-Arias A, et al. The estimation of the effective reproductive number from disease outbreak data. Math Biosci Eng. 2009;6(2):261–82.

9. Ferguson NM, Donnelly CA, Anderson RM. Transmission intensity and impact of control policies on the foot and mouth epidemic in Great Britain. Nature. oct 2001;413(6855):542–8.

10. White LF, Pagano M. Transmissibility of the influenza virus in the 1918 pandemic. PloS One. 30 janv 2008;3(1):e1498.

11. Cauchemez S, Boëlle PY, Thomas G, Valleron AJ. Estimating in real time the efficacy of measures to control emerging communicable diseases. Am J Epidemiol. 15 sept 2006;164(6):591–7.

12. Cori A, Boëlle PY, Thomas G, Leung GM, Valleron AJ. Temporal variability and social heterogeneity in disease transmission: the case of SARS in Hong Kong. PLoS Comput Biol. août 2009;5(8):e1000471.

13. Hens N, Van Ranst M, Aerts M, Robesyn E, Van Damme P, Beutels P. Estimating the effective reproduction number for pandemic influenza from notification data made publicly available in real time: a multi-country analysis for influenza A/H1N1v 2009. Vaccine. 29 janv 2011;29(5):896–904.

14. Cori A, Ferguson NM, Fraser C, Cauchemez S. A New Framework and Software to Estimate Time-Varying Reproduction Numbers During Epidemics. Am J Epidemiol. 1 nov 2013;178(9):1505–12.

15. White LF, Moser CB, Thompson RN, Pagano M. Statistical Estimation of the Reproductive Number From Case Notification Data. Am J Epidemiol. 6 avr 2021;190(4):611–20.

16. Amundsen EJ, Stigum H, Røttingen JA, Aalen OO. Definition and estimation of an actual reproduction number describing past infectious disease transmission: application to HIV epidemics among homosexual men in Denmark, Norway and Sweden. Epidemiol Infect. éc 2004;132(6):1139–49.

17. Bettencourt LMA, Ribeiro RM. Real Time Bayesian Estimation of the Epidemic Potential of Emerging Infectious Diseases. PLOS ONE. 14 mai 2008;3(5):e2185.

18. Cintrón-Arias A, Castillo-Chávez C, Bettencourt LMA, Lloyd AL, Banks HT. The estimation of the effective reproductive number from disease outbreak data. Math Biosci Eng MBE. avr 2009;6(2):261–82.

19. Ferguson NM, Donnelly CA, Anderson RM. Transmission intensity and impact of control policies on the foot and mouth epidemic in Great Britain. Nature. 4 oct 2001;413(6855):542–8.

20. Ferrari MJ, Bjørnstad ON, Dobson AP. Estimation and inference of R0 of an infectious pathogen by a removal method. Math Biosci. nov 2005;198(1):14–26.

21. Forsberg White L, Pagano M. A likelihood-based method for real-time estimation of the serial interval and reproductive number of an epidemic. Stat Med. 2008;27(16):2999–3016.

22. Fraser C, Donnelly CA, Cauchemez S, Hanage WP, Van Kerkhove MD, Hollingsworth TD, et al. Pandemic potential of a strain of influenza A (H1N1): early findings. Science. 19 juin 2009;324(5934):1557–61.

23. Griffin JT, Garske T, Ghani AC, Clarke PS. Joint estimation of the basic reproduction number and generation time parameters for infectious disease outbreaks. Biostat Oxf Engl. avr 2011;12(2):303–12.

24. Howard SC, Donnelly CA. Estimation of a time-varying force of infection and basic reproduction number with application to an outbreak of classical swine fever. J Epidemiol Biostat. 2000;5(3):161–8.

25. Kelly HA, Mercer GN, Fielding JE, Dowse GK, Glass K, Carcione D, et al. Pandemic (H1N1) 2009 influenza community transmission was established in one Australian state when the virus was first identified in North America. PloS One. 28 juin 2010;5(6):e11341.

26. Riley S, Fraser C, Donnelly CA, Ghani AC, Abu-Raddad LJ, Hedley AJ, et al. Transmission dynamics of the etiological agent of SARS in Hong Kong: impact of public health interventions. Science. 20 juin 2003;300(5627):1961–6.

27. Wallinga J, Lipsitch M. How generation intervals shape the relationship between growth rates and reproductive numbers. Proc R Soc B Biol Sci. 22 févr 2007;274(1609):599–604.

28. Wallinga J, Teunis P. Different Epidemic Curves for Severe Acute Respiratory Syndrome Reveal Similar Impacts of Control Measures. Am J Epidemiol. 15 sept 2004;160(6):509–16.

29. Cauchemez S, Boelle PY, Donnelly CA, Ferguson NM, Thomas G, Leung GM, et al. Real-time estimates in early detection of SARS. Emerg Infect Dis. janv 2006;12(1):110–3.

30. Obadia T, Haneef R, Boëlle PY. The R0 package: a toolbox to estimate reproduction numbers for epidemic outbreaks. BMC Med Inform Decis Mak. 18 déc 2012;12(1):147.

31. Wood SN. Generalized Additive Models: An Introduction with R. CRC Press; 2017.

32. Craven P, Wahba G. Smoothing noisy data with spline functions. Numer Math. 1 éc 1978;31(4):377–403.

33. Burton A, Altman DG, Royston P, Holder RL. The design of simulation studies in medical statistics. Stat Med. 30 déc 2006;25(24):4279–92.

34. Morris TP, White IR, Crowther MJ. Using simulation studies to evaluate statistical methods. Stat Med. 20 mai 2019;38(11):2074–102.

35. Guerra FM, Bolotin S, Lim G, Heffernan J, Deeks SL, Li Y, et al. The basic reproduction number (R0) of measles: a systematic review. Lancet Infect Dis. déc 2017;17(12):e420–8.

36. Organisation mondiale de la santé. Rougeole [Internet]. [cité 28 avr 2023]. Disponible sur: https://www.who.int/fr/news-room/fact-sheets/detail/measles

37. European Centre for Disease Prevention and Control. Factsheet about measles [Internet]. 2017 [cité 28 avr 2023]. Disponible sur: https://www.ecdc.europa.eu/en/measles/facts

38. Epicentre - Médecins sans frontières (MSF). Rougeole [Internet]. [cité 3 juill 2023]. Disponible sur: https://epicentre.msf.org/nos-concretisations/rougeole

39. Cori A,, Cauchemez S, Ferguson NM, Fraser C, Dahlqwist E, et al. EpiEstim: Estimate Time Varying Reproduction Numbers from Epidemic Curves [Internet]. 2021 [cité 5 août 2024]. Disponible sur: https://cran.r-project.org/web/packages/EpiEstim/index.html

40. Gostic KM, McGough L, Baskerville EB, Abbott S, Joshi K, Tedijanto C, et al. Practical considerations for measuring the effective reproductive number, Rt. PLoS Comput Biol. 10 éc 2020;16(12):e1008409.

41. Hastie TJ, Tibshirani RJ. Generalized Additive Models. CRC Press; 1990. 356 p.

42. Meyer S, Held L, Höhle M. Spatio-Temporal Analysis of Epidemic Phenomena Using the R Package surveillance. J Stat Softw. 3 mai 2017;77:1–55.

43. Rigby RA, Stasinopoulos DM. Generalized additive models for location, scale and shape. J R Stat Soc Ser C. 2005;54(3):507–54.

